# Use of unstructured text in prognostic clinical prediction models: a systematic review

**DOI:** 10.1101/2022.01.17.22269400

**Authors:** Tom M. Seinen, Egill Fridgeirsson, Solomon Ioannou, Daniel Jeannetot, Luis H. John, Jan A. Kors, Aniek F. Markus, Victor Pera, Alexandros Rekkas, Ross D. Williams, Cynthia Yang, Erik van Mulligen, Peter R. Rijnbeek

## Abstract

**Objective:** This systematic review aims to assess how information from unstructured clinical text is used to develop and validate prognostic risk prediction models. We summarize the prediction problems and methodological landscape and assess whether using unstructured clinical text data in addition to more commonly used structured data improves the prediction performance.

**Materials and Methods:** We searched Embase, MEDLINE, Web of Science, and Google Scholar to identify studies that developed prognostic risk prediction models using unstructured clinical text data published in the period from January 2005 to March 2021. Data items were extracted, analyzed, and a meta-analysis of the model performance was carried out to assess the added value of text to structured-data models.

**Results:** We identified 126 studies that described 145 clinical prediction problems. Combining text and structured data improved model performance, compared to using only text or only structured data. In these studies, a wide variety of dense and sparse numeric text representations were combined with both deep learning and more traditional machine learning methods. External validation, public availability, and explainability of the developed models was limited.

**Conclusion:** Overall, the use of unstructured clinical text data in the development of prognostic prediction models has been found beneficial in addition to structured data in most studies. The EHR text data is a source of valuable information for prediction model development and should not be neglected. We suggest a future focus on explainability and external validation of the developed models, promoting robust and trustworthy prediction models in clinical practice.

## INTRODUCTION

Prognostic prediction models are increasingly common in clinical research and practice [1,2]. A prognostic model predicts which patients, among a target population of patients, will experience some clinical outcome during a prediction horizon, using predictors measured during an observation window prior to the time of prediction. The growing availability of observational data in electronic health records (EHRs) forms a rich source to develop prediction models in a data-driven manner [2,3]. Although most clinical risk prediction research is centered on the use of structured EHR data, such as coded conditions, measurements, and drug prescriptions, the majority of information in EHRs is typically stored in vast quantities of unstructured text, for example, nursing notes, discharge letters, or radiology reports [4]. Compared to structured data, clinical free text lacks an organized structure or terminology, is large in terms of file size, and contains patient-sensitive information, which complicates its use for the construction of prediction models. However, information captured in text can be more detailed and extensive than in the structured data, as it is not limited to specific code systems or input fields. Therefore, incorporating this information could potentially improve prediction model performance.

The growing availability of unstructured text in EHR data, increased computational power, and progress in natural language processing (NLP) techniques are now enabling the use of text data for the development of prediction models. Textual data has already been used in the clinical domain for a variety of tasks. Several reviews have elucidated the adoption of clinical NLP, focusing primarily on the general task of extracting information from clinical notes [5-11] or diagnostic classification of patients, including case detection, patient identification, and phenotyping [4,9,12]. A recent systematic review by Yan et al. [13] studied the use of clinical text in early sepsis prediction. However, to our knowledge, no broad systematic review has been conducted on the development of prognostic prediction models incorporating unstructured clinical text. As text-based prognostic prediction models start to be developed, it becomes increasingly important to reflect on the work that has been done, summarize the methodological landscape, and discover whether text data has value supplementing coded data.

Consequently, the objective of this review is to assess how information extracted from unstructured EHR text data is utilized to develop and validate prognostic prediction models. We evaluated the studies on the study settings and populations, text processing methods and representations, machine-learning methods and feature sets combinations, performance evaluation and external validation, and model explainability and availability. Furthermore, we determined the value of text in addition to structured data by comparing the performance between models using these different feature sets within the studies.

## MATERIALS AND METHODS

### Review protocol

The review protocol for this study was registered on June 17, 2021, and is publicly available at the Open Science Framework Registries (https://osf.io/gw628).

### Eligibility criteria

This review targeted studies from the last 15 years (January 2005 to March 2021) describing the development and evaluation of prognostic clinical prediction models that incorporate information extracted in a data-driven manner from unstructured EHR text data. The three inclusion criteria are defined as follows. (1) The study described the development and evaluation of a prognostic clinical prediction model. (2) The model predictors were based on information extracted from unstructured EHR text data. (3) Information was automatically extracted from the unstructured text in a data-driven manner. Data-driven implies that the extraction of information from the text was exploratory and not restricted to features that were expected to be important. An extensive description of the inclusion criteria is provided in the supplementary material.

### Literature search

Four databases were used for the literature search: Embase, MEDLINE, the Web of Science core collection, and Google Scholar. The database choice and the search strategy creation was aided by a medical librarian. The search strategy consisted of four clauses that incrementally limited the search results: (1) Prediction models; (2) The medical domain or electronic health records; (3) A notion of text data, clinical notes, or NLP methods; (4) The period from January 2005 till March 2021, studies in the English language, and excluding conference abstracts and animal research. The full search strategy can be found in Table S1.

### Screening

All studies found in the literature search were first screened for fulfilling the eligibility criteria based on the title and abstract. Those that were found relevant underwent a second screening for inclusion based on the full text. In both screening phases, one reviewer (TS) screened all studies and ten other reviewers (EF, SI, DJ, LJ, JK, AM, VP, AR, RW, CY) independently screened one-tenth of the total number of studies. This resulted in each study being screened by two independent reviewers. Any discrepancies between them, in both screening phases, were resolved in a consensus meeting.

### Data extraction and synthesis

Data were extracted from the included studies by one reviewer (TS) using a pre-defined set of data items. The set is outlined in Table 1, including the type of input. Some items are based on clinical prediction item sets from the *critical appraisal and data extraction for systematic reviews of prediction modelling studies* (CHARMS) checklist [14] and the *transparent reporting of a multivariable prediction model for individual prognosis or diagnosis* (TRIPOD) statement [15]. Ten data item topics were distinguished: (1) the general publication information, (2) the study setting, (3) the study population, (4) unstructured text data predictors, (5) structured data predictors, (6) machine-learning methods and feature sets, (7) the internal and (8) external validation, (9) model explainability, and (10) model availability.

**Table 1.** List of data items for data extraction, by topic. Data item sources indicated by A: CHARMS and B: TRIPOD; an asterisk (*) indicates data items added to the review protocol.

| Item topic | Data item | Input type |
| --- | --- | --- |
| 1. General information | Publication year | Year |
|  | Journal | Free text |
| 2. Study setting | Dataset | Free text |
|  | Country of data | Country |
|  | Clinical setting | Free text |
|  | Study dates | Range of years |
| 3. Population | Type of study* | Cohort, Case-control |
|  | Target population<sup>AB</sup> | Free text |
|  | Prediction outcome<sup>A</sup> | Free text |
|  | Prediction time horizon<sup>A</sup> | Hours, Days, Years, Relative time, Timepoint |
|  | Prediction outcome type | Binary, Multiclass, Continues |
| 4. Unstructured text data predictors | Type of text content | Free text |
|  | Language of text | Language |
|  | Observation window<sup>B</sup> | Hours, Days, Years, Relative time, Timepoint |
|  | Pre-processing methods | Free text |
|  | Text representation methods | Bag-of-Words (BoW), TFIDF, Concept Extraction (CE), Word Embedding (WE), Document Embedding (DE), Topic model (TM), Summarizing Score (SS). (multiple possible) |
|  | Used ontologies/vocabularies | Free text |
|  | Used software/program/package | Free text |
|  | Number of predictors<sup>A</sup> | Number |
| 5. Structured data predictors | Types of structured data | Free text |
|  | Observation window<sup>B</sup> | Hours/Days/Years/Relative time/Timepoint |
|  | Number of predictors<sup>A</sup> | Number |
| 6. Model | Machine-learning method<sup>A</sup> | Logistic regression (LogR), linear regression (LinR), cox proportional hazards regression (Cox), Naïve Bayes (NB), Random forests or other tree based methods (RFTB), Gradient Boosting (GB), Support Vector Machines (SVM), (simple) Neural Networks (NN), Recurrent Neural Networks (RNN), Convolutional Neural Networks (CNN), Transformer, (other) Deep Neural Networks (DNN), Ensembles, Other. |
|  | Feature-set | Structured/Text/Combined |
| 7. Internal validation | Number of subjects/observations<sup>A</sup> | Number |
|  | Number of cases<sup>A</sup> | Number |
|  | AUC, AUPRC, F1-score<sup>A</sup> | Values |
|  | Accuracy, sensitivity (or recall), specificity, and positive predictive value (or precision) reported?<sup>A</sup> | Yes/No |
|  | MSE/MAE reported?<sup>A</sup> | Yes/No |
|  | ROC/PR curves presented?<sup>A</sup> | Yes/No |
|  | Calibration plot or metrics presented?<sup>AB</sup> | Yes/No |
| 8. External validation | Type of external validation<sup>A</sup> | Same/another department/center/country |
|  | Same items as internal validation |  |
| 9. Explainability | Global feature importance presented?* | Yes/No |
|  | Single patient (local) feature importance presented?* | Yes/No |
| 10. Final model availability | Is the final model directly available to apply to different data?<sup>A</sup> | Yes/No |
|  | Is the study code available to reproduce the methods?* | Yes/No |

The input for text representation methods consisted of a list of both sparse and dense numeric vector representations. Sparse representations included Bag-of-Words (*BoW*), a Bag-of-Words variant: Term Frequency – Inverse Document Frequency (*TFIDF*), and clinical concept extraction (*CE*). Dense vector representations included topic models (*TM*), word and document embeddings (*WE, DE*), and summarizing scores (*SS*), such as a sentiment score. Combinations of representations were possible. The machine learning methods were of varying complexity and interpretability, including methods ranging from simple linear or logistic regression (*LinR, LogR*), random forests or other tree-based methods (*RFTB*), gradient boosting (*GB*), and Support Vector Machines (*SVM*), to complex deep neural networks (*DNN*).

If a study reported on multiple prediction problems, the data items were extracted for each reported problem. The model and validation data items were only extracted for the – self-reported – best performing structured-data, text-data, and combined-data models in each problem. For data items with free text input, the results were manually categorized after data extraction to enable analysis. We performed a meta-analysis on the reported model performance, comparing the structured-data, text-data, and combined-data models, for each prediction problem. The differences in the area under the receiver operator curve (AUC) metric were calculated for each reported feature-set comparison: text and structured data (ΔAUC_TS_ = AUC_T_ - AUC_S_), combined and structured data (ΔAUC_CS_ = AUC_C_ - AUC_S_), and combined and text data (ΔAUC_CT_ = AUC_C_ - AUC_T_). The AUC differences indicate the relative performance difference between the use of the three feature sets within each prediction problem and are suitable to be compared across studies.

## RESULTS

### Search and data-extraction results

The literature search, performed in March 2021, resulted in a total of 5043 studies. The PRISMA flow diagram is presented in Figure 1. After deduplication, removing 2030 studies, a set of 3013 studies was screened on title and abstract. We excluded 2783 studies that violated one of the inclusion criteria. Full-text screening of the remaining 230 studies resulted in 126 relevant studies to be included in the review. The 104 studies that were excluded based on their full text consisted of 5 duplicate studies, 52 studies not performing prognostic modelling or a performance evaluation, 13 studies with no use of text data in the prediction model, 28 studies without data-driven information extraction, and 6 studies with other reasons for exclusion (no full-text available, not peer-reviewed, reviews).

**Figure 1.**
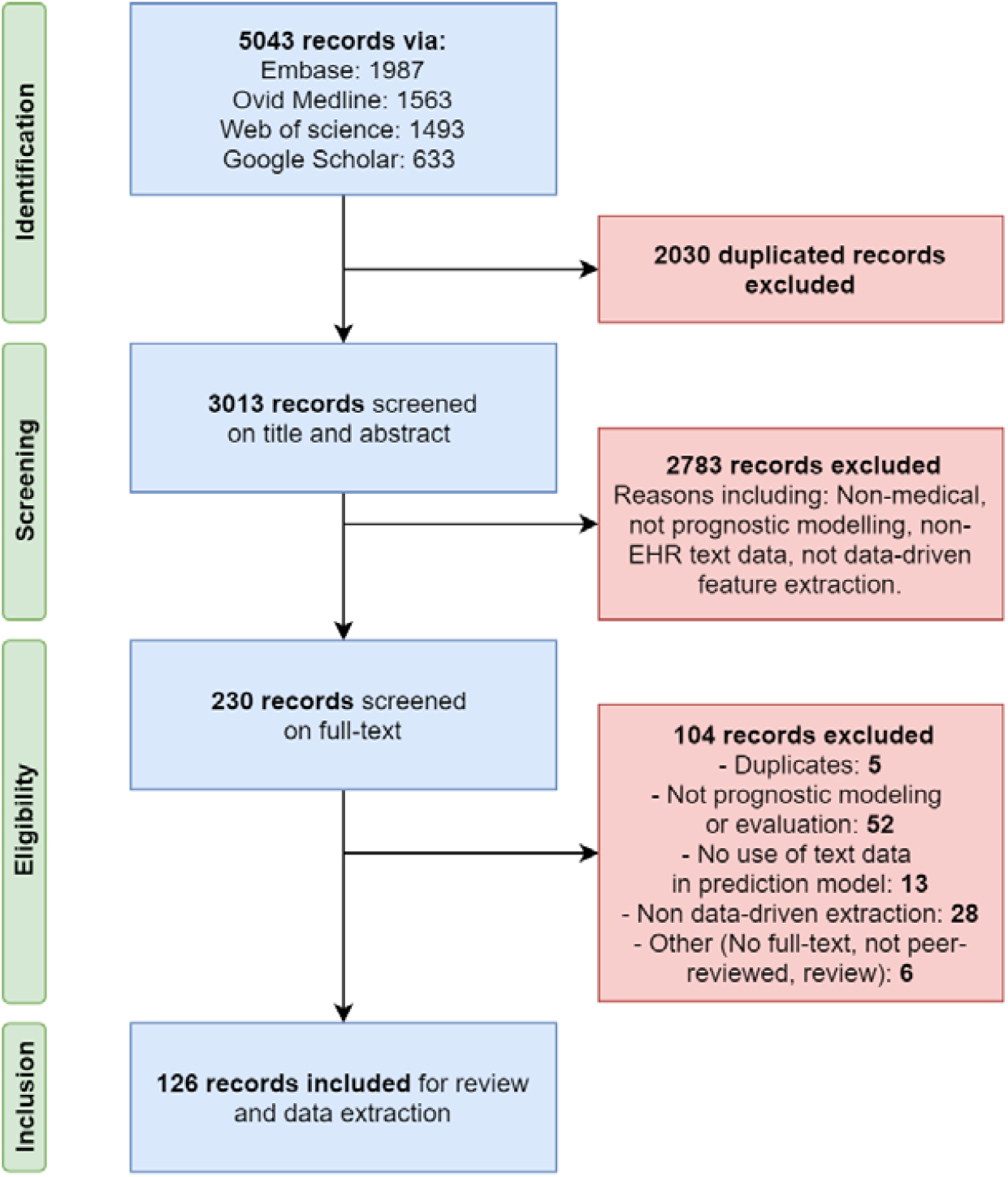
PRISMA flow diagram with the search and screening results of the systematic review.

We extracted the different data items for each study and its reported prediction problems. Fourteen of the 126 studies reported multiple prediction problems, resulting in a total of 145 prediction problems. A list of characteristics of the studies ordered by publication year is presented in Table S2 and the extracted data items are available as supplementary data. The large majority of the reviewed studies (79%) was published in the period from January 2018 to March 2021 (Table 2). No eligible studies were found in the period 2005 to 2011. The studies were published in a variety of journals and conference proceedings. The journals with the highest number of studies were the Journal of Biomedical Informatics (9%), PLoS ONE (6%), BMC Medical Informatics and Decision Making (5%), JMIR Medical Informatics (5%), and the Journal of the American Medical Informatics Association (5%).

**Table 2.** Number of included studies by publication year.

| Year | 2012 | 2013 | 2014 | 2015 | 2016 | 2017 | 2018 | 2019 | 2020 | 2021<br>(until March) |
| --- | --- | --- | --- | --- | --- | --- | --- | --- | --- | --- |
| Number of included studies | 4 | 0 | 5 | 4 | 9 | 5 | 19 | 30 | 41 | 9 |

Around half of the reviewed studies compared models that used structured data (S), text data (T), or a combination of structured and text data (C). A comparison between all three feature sets (S:T:C) was reported for 23% of the prediction problems. In 27% of the problems two feature sets were compared (S:T 4%; S:C 19%; T:C 3%) and in 50% no comparison was made and the use of only one feature set was reported (T 33%; C 17%).

### Clinical settings and prediction problems

Most prediction problems focused on general hospital care settings (47%), followed by intensive care (18%), emergency care (14%), surgical care (8%), and psychiatric or mental health care (7%). Only a few prediction problems (6%) were set in outpatient, or radiology settings. Almost half (47%) of the prediction problems used a local proprietary dataset, 13% used a collection of two or more local datasets, and 7% used registry, claims, or survey datasets. One third (33%) of the prediction problems was developed on a publicly available dataset, specifically, in 47 out of 48 cases a *Medical Information Mart for Intensive Care* (MIMIC-II/III) database [16,17] was used.

Figure 2 visualizes the different categories of target populations, clinical outcomes, and prediction horizons that make up each prediction problem. The three largest target populations were patients with general hospital admissions (22%), intensive care (ICU) admissions (18%), and emergency department (ED) visits (14%). The largest outcome events were mortality (29%), diagnosis of a specific disease or condition (19%), and hospital or ICU readmission (12%). Most prediction problems had as prediction horizon a period of months (30%) or the period of hospital/ICU admission (27%). There were 82 unique combinations of which 58 only occurred once. The prediction of mortality during admission in ICU patients occurred most often (7%), followed by the prediction of admission or transfer at ED discharge for patients visiting the ED (6%). The observation window prior to the time of prediction was not reported in 15% of the problems. The most-reported observation windows were the first hours of a hospital or ICU admission or ED visit (20%), the entire time of admission or the visit (15%), and during triage (10%).

**Figure 2.**
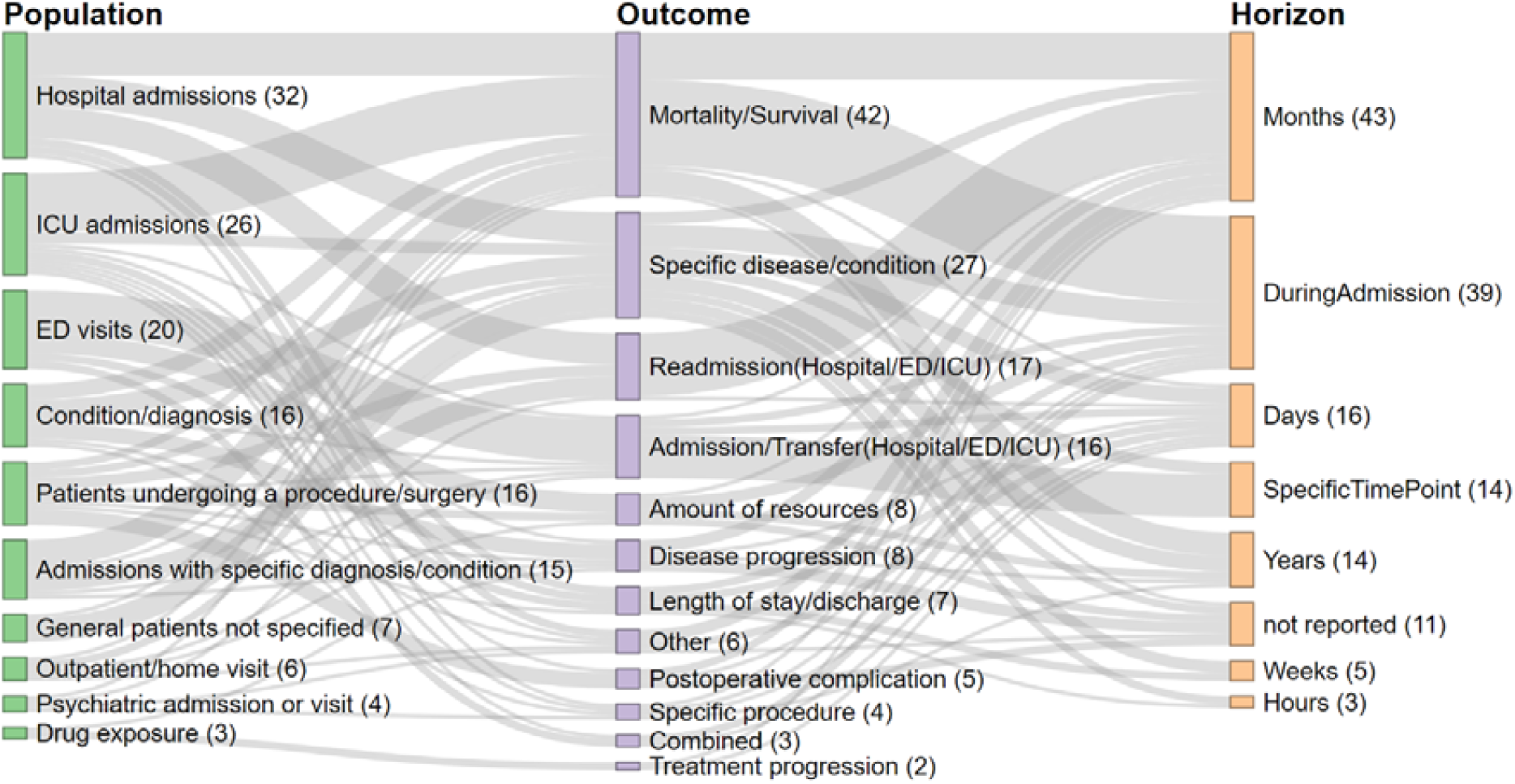
Sankey diagram of the different categories of target populations and clinical outcomes, and clinical outcomes and prediction horizons, ordered by size. The number in parentheses indicates the number of prediction problems with these categories and the width of the connection between two categories represents the number of prediction problems with this combination of categories.

The distribution of the number of observations and outcome cases is depicted in Figure 3A together with the distribution of their ratio in Figure 3B. The number of observations differed much between studies, from only a few hundred observations to a few million, with an mean of 87,016 and a median of 17,973 observations. Observations and outcome cases had an average ratio of 0.20 and a median of 0.14. In only 0.2% of prediction problems, the number of observations was not reported and in 16% the number of outcome cases was missing.

**Figure 3.**
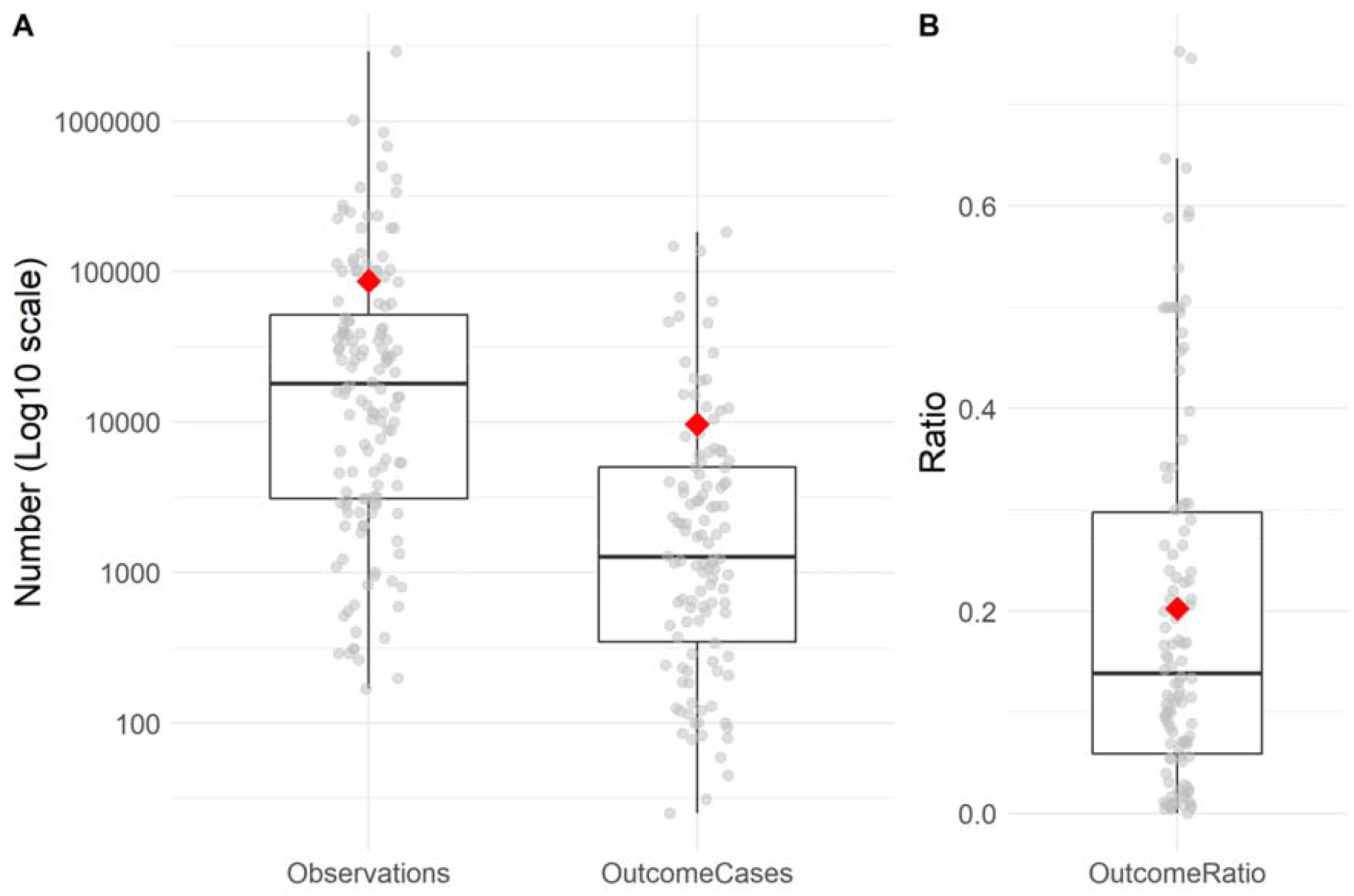
A: Boxplots of the number of observations (left) and outcome cases (right) of 145 prediction problems. B: Boxplot of the ratios between the number of observations and outcome cases. In both A and B the mean is indicated by the red diamond and the grey points represent the underlying data.

### Pre-processing methods and text representations

Text pre-processing methods, applied before the text representation creation, were well-reported in 74% of prediction problems. The pre-processing of text commonly included methods such as sentence splitting, tokenization, lemmatization, the removal of stop words, punctuation, or numbers, abbreviation disambiguation, and the filtering of tokens based on frequencies. The text data was written in English for the majority (79%) of the prediction problems, followed by Chinese (6%) and Portuguese (3%). In total 12 different languages were reported.

Bag-of-Words text representations (including TFIDF) were used most often in 36% of the text and combined-data models, followed by word embeddings (18%) and concept extraction (14%). In some cases, multiple representations were combined by concatenation (*Combination*) (6%) or dense representation were generated from extracted concepts (*CE/WE, CE/TM*) (3%). Dense representations had a median dimension of 200 features against 6985 features in sparse representations (Figure S1). Pre-processing methods were more frequently reported together with the use of Bag-of-Words (85%) and TFIDF (92%) compared to concept extraction (54%), document embeddings (58%), and word embedding (75%) methods, which often used out-of-the-box tools or software. *MetaMap* [18] was the most common software used for extracting clinical concepts from text data, in 11 out of the 30 models using extracted concepts. Concept extraction also requires a clinical ontology or vocabulary as a reference for coded concepts. The full set or a part of the UMLS vocabularies [19] was used most, for 26 models. Five models made only use of the SNOMED Clinical Terms [20] and for another four no ontology was reported.

### Machine learning methods

The most used methods for training text and combined-data models were logistic regression (27%), recurrent neural networks (13%), and random forest or other tree-based methods (10%). For the structured-data models, logistic regression was also the most prevalent method (30%), followed by gradient boosting (20%) and recurrent neural networks (12%). In the large majority (89%) of prediction problems, a binary prediction model was trained, followed by multi-class prediction (7%) and the prediction of a continuous outcome (4%).

Figure 4A depicts the use of both text representations (abstracted as dense, sparse, or combined representations) and machine-learning methods (traditional, (deep) neural networks, or ensemble methods) in both text and combined-data models over the years. It can be observed that until 2017 the use of sparse representations and traditional models was most common, but after 2018 the use of both dense text representations and (deep) neural networks took off. To understand their joined rise we examined the relationship between the model’s machine-learning method and its textual input representation in Figure 4B. It shows that the dense word and document embeddings served primarily as the input for deep learning methods, while the Bag-of-Words representations were commonly used by more traditional machine learning methods. The text representations and machine-learning methods are significantly associated, X (4, N=183)=36.1, p<.001.

**Figure 4.**
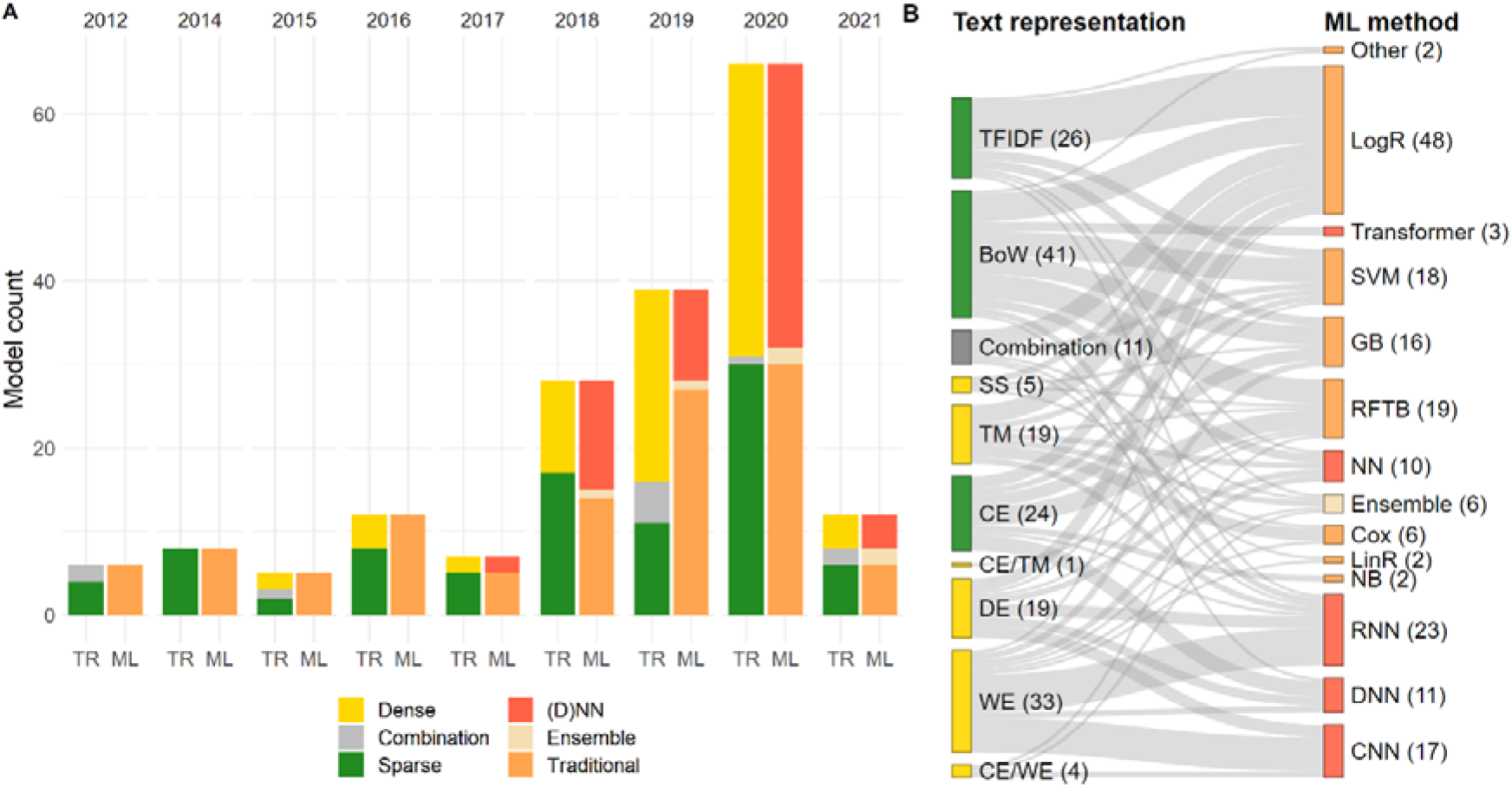
A: The use of different text representations (TR) and machine-learning (ML) methods in text-based or combined-data prediction models over time. No eligible studies in 2013. B: The combinations of text representations (left) and machine-learning methods (right) in text-based or combined-data prediction models. The number in parentheses indicates the number of prediction problems with these categories and the width of the connection between two categories represents the number of prediction problems with this combination of categories. Abbreviations can be found in Table 1.

### Model performance evaluation and comparison

The internal validation model performance was reported using the AUC (or c-statistic/index) for the majority (83%) of prediction models. For the other models only metrics that are based on dichotomized outcomes, such as accuracy, sensitivity (or recall), specificity, and positive predictive value (or precision), were reported. The mean squared error or mean absolute error were reported for regression models predicting continuous outcomes. The F1-score was reported for 31% of the models and the area under the precision-recall curve (AUPRC) for 14%. The combined reporting of metrics is visualized in Figure S2. A receiver operator curve or precision-recall curve was presented for 39% of problems, but for only 12% a calibration plot or calibration metric (such as the brier-score or calibration intercept and slope) was presented.

Figure 5A depicts the distributions of AUC differences (ΔAUC) between the structured-data, text-data, and combined-data models within each prediction problem. The combined-data models had a visibly higher performance than the text or structured-data models and the average AUC differences, for both ΔAUC_CS_ and ΔAUC_CT_, were significantly larger than zero, *t*(53)=6.76, *p*<.001 and *t*(33)=5.49, *p*<.001 respectively. Text-data models did not perform significantly better or worse than the structured-data models. Their AUC difference, ΔAUC_TS_, showed a large variation across prediction problems. We investigated whether there was a relationship between these AUC differences and the clinical settings. Figure 5B shows how the text and structured-data model performance differences vary between four clinical settings: emergency, hospital, intensive, and surgical care. Psychiatric care is not presented as it only had one observation. Following a full pairwise comparison of the distributions, we found that the AUC difference means of the intensive and surgery care prediction problems were different *t*(8.55)=-3.95, *p*=.024 (Bonferroni adjusted). This implies that models using text data in the surgical care setting had on average a higher performance (compared to structured data models) than in the intensive care setting.

**Figure 5.**
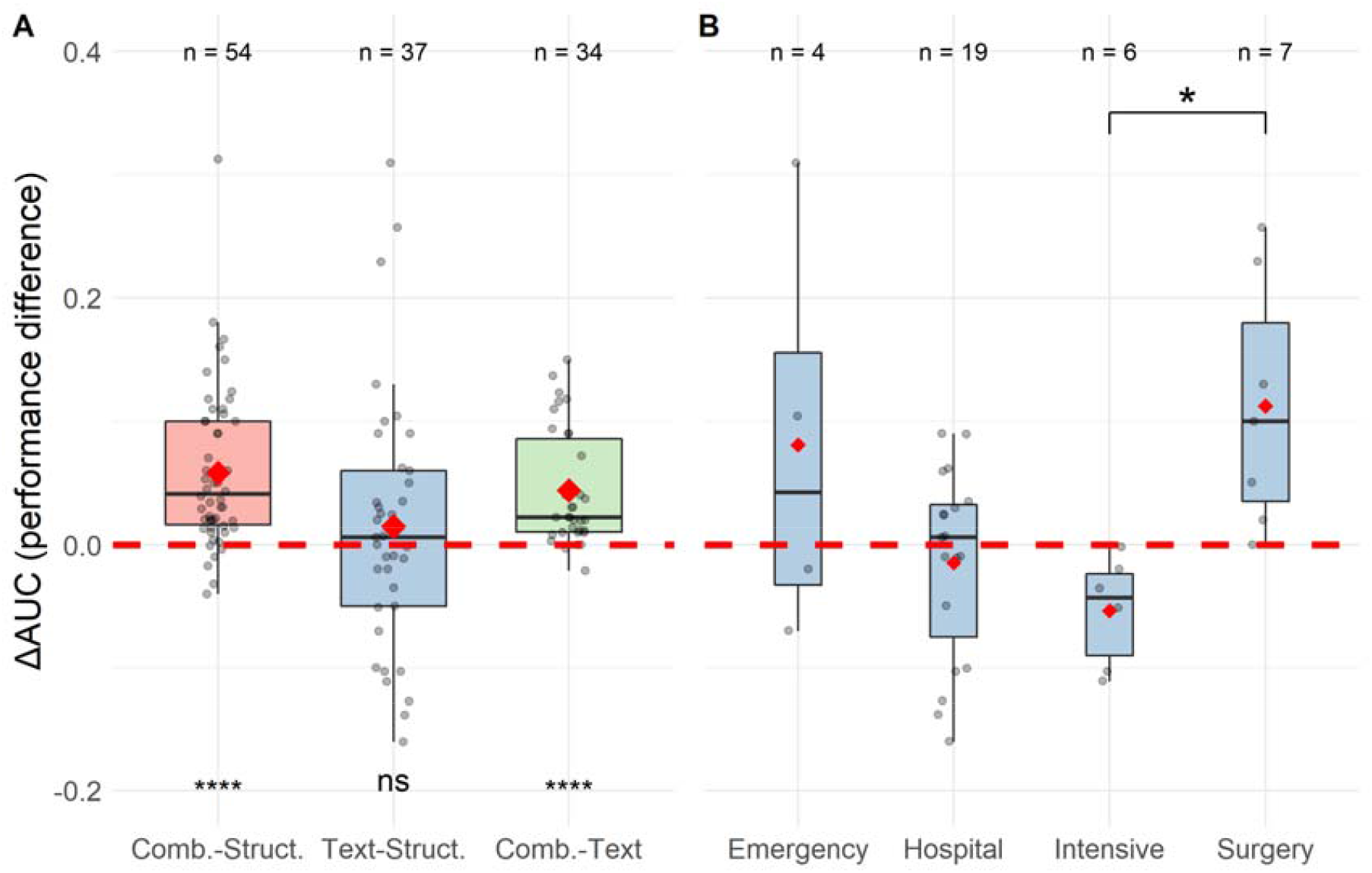
A: AUC difference distribution boxplots of the combined and structured-data models (ΔAUC_CS_), the text and structured-data models (ΔAUC_TS_), and combined and text-data models (ΔAUC_CT_). B: Text and structured-data model AUC difference (ΔAUC_TS_) boxplots for four different clinical settings. In both A and B, the means are indicated by a red diamond, the grey points represent the underlying data, sample sizes are shown on top, and the red dotted line indicates the AUC difference of zero. ns=not significant, *=p<.05, ****=p<.001.

Notably, prediction models were externally validated in only two studies. Marafino et al. [21] externally validated their in-hospital mortality prediction model within three medical centers and Menger et al. [22] externally validated their in-hospital patient violence prediction model at two sites. Both studies reported a small to moderate decrease in external validation model performance, between 0.023 and 0.079 AUC difference.

### Model explainability and availability

Global feature importance, the feature importance over all predictions, was reported for 44% of the prediction problems and a local feature importance, the contribution of features for a specific prediction, was presented in 6% of problems. The final model was presented or made available in only 5% of prediction problems, while the code used for training the model was in 21% directly available online.

## DISCUSSION

### Model performance

We found that in the 126 studies developing prognostic clinical prediction models using unstructured EHR text data, published in the last 15 years, the performance of combined-data models on average outperformed the text and structured-data models. This demonstrates that the available EHR text data is a source of valuable information able to improve model performance in addition to structured data. Yan et al. [13] found comparable results in a review of nine studies predicting sepsis. While on average text-data models did not outperform structured-data models, we did see an interesting difference between clinical settings. In intensive care prediction problems, the structured-data models had a higher performance than the text-data models when compared to surgical care. This may be explained by the inherent differences in clinical documentation and code registration between the clinical settings. Intensive care is generally a structured-data rich setting, while in surgical care the information is contained in surgical reports. It shows that the distribution of information over text and structured data is likely dependent on various factors.

### Clinical settings, datasets, and language

Hospital care (including intensive, surgical, and emergency care) was the most common clinical setting. While the combinations of different target populations, outcome events, and prediction horizons varied much between prediction problems, common themes could be observed, such as the prediction of mortality in the ICU or ED discharge disposition. Almost half of the studies used a proprietary dataset and most studies using public datasets (almost a third of the reviewed studies) relied on a MIMIC dataset [16]. Being one of the few large public clinical datasets containing anonymized unstructured data, MIMIC enables transparent and reproducible research on clinical text and serves as a benchmark for clinical NLP tasks. Almost 90% of the reviewed studies were performed on English clinical text. This suggests that opportunities still exist for studying model development using unstructured text in other languages [23,24].

### Text processing and machine learning methods

The techniques used for pre-processing text and creating numeric text representations were generally well described. The impact of pre-processing methods on the model performance can be significant and those methods are therefore essential to report [25]. The sparse Bag-of-Words and TFIDF representations and the dense word and document embeddings were most frequently used and we found an association between the types of text representation and machine learning methods. The (deep) neural network methods generally used a dense text representation, while regularized logistic regression methods, random forests, or SVMs largely took sparse representations as input.

### Model explainability and External validation

Global or local feature importance was presented for less than half of the prediction problems, which may be considered rather limited given the importance of model explainability and trustworthiness in clinical risk prediction [26]. Compared to simple logistic regression models, deep learning models need additional effort to be explained. Deep learning is well-suited for handling and combining structured and unstructured data [27], but the high complexity and dense input features impede direct explainability without post-hoc explanation techniques [26].

Furthermore, only two studies presented external validation results and relatively few studies shared their trained model or code. Externally validating prediction models using text data might be challenging, due to the differences in (sub)language and EHR systems or the fear of sharing identifiable patient information captured by the model. However, assessing generalizability and external validity remains important in model development [28]. Frameworks exist, such as the Observational Medical Outcomes Partnership Common Data Model (OMOP CDM) [29], that deal with the lack of syntactic and semantic interoperability in health data. The OMOP CDM enables external validation by evaluating trained prediction models on other databases, only reporting back the aggregated and anonymized results, and it allows research to meet the challenges of validating text-based models between databases using different languages [1,3].

We suggest a future research focus on model explainability and external validation, and advocate the sharing of code or trained models for external validation by others. These steps will not only expand the model’s generalizability but will also promote the use of robust and trustworthy prediction models in clinical practice [28].

### Strengths and limitations

There were likely some published studies eligible for inclusion that we did not find. For example, studies that incorporated text data in a prediction model but did not mention it in the title or abstract would have been missed. Nonetheless, the search query was designed to capture a wide variety of terms that would indicate the use of unstructured text data. Furthermore, the level of granularity for predefined categories for text representations or machine-learning methods was high. More granular categories on, for example, the different deep learning architectures could have been collected for more detailed and in-depth information. However, this would also have resulted in decreasing numbers per category, complicating interpretation. To the best of our knowledge, this is the first systematic review on the development of prognostic clinical prediction models using unstructured text. We performed a broad literature search over a long period of time, resulting in a large set of eligible studies in a wide variety of clinical settings, not focused on one specific prediction problem. The comparison of the relative performance between text, structured, and combined feature sets within each study allowed us to assess the value of text data in prediction model development. Finally, we made the extracted data available to provide transparency and reproducibility.

## CONCLUSION

In this systematic review, we found that the use of unstructured clinical text data in the development of prognostic prediction models was beneficial in most of the studies. Combining unstructured text with structured data in prognostic prediction model development generally improved model performance, while the performance of text-data models compared to structured-data models varied. Overall, unstructured text in EHR data should not be neglected, as it is a source of valuable information that can improve prediction model performance in addition to structured data. But the information available in both structured and unstructured data is likely dependent on multiple factors, such as the clinical setting. Models were generally developed in hospital care settings using a variety of text representations and machine-learning methods and we found a relationship between the types of text representation and machine-learning methods used. Furthermore, it is a cause for concern that only two studies externally validated their developed prediction models and that many studies had little attention for model explainability. Therefore, we suggest a focus on external validation and model explainability in future research. Additionally, we emphasize the importance of studying the use of text in non-English languages in prediction model development.

## Data Availability

The data underlying this work are available as supplementary material.

## FUNDING

This work has received support from the European Health Data & Evidence Network (EHDEN) project. EHDEN has received funding from the Innovative Medicines Initiative 2 Joint Undertaking (JU) under grant agreement No 806968. The JU receives support from the European Union’s Horizon 2020 research and innovation programme and EFPIA.

ACKNOWLEDGEMENTS

The authors would like to thank Christa Niehot from the Erasmus MC medical library for her input on the search strategy.

## DATA AVAILABILITY STATEMENT

The extracted data used for generating the results are available as supplementary data.

