## Supplementary material, Table S1, Table S2, Figure S1, Figure S2 for "Use of unstructured text in prognostic clinical prediction models: a systematic review"

### **Inclusion criteria**

1. **The study must describe the development and evaluation of a prognostic clinical risk prediction model.**
   a. This review is focused on prognostic prediction, i.e. predicting a future clinical event or outcome for a patient.
      - Studies that develop a diagnostic, identification, phenotyping, or extraction model are excluded.
   b. The subject must be a patient or a limited group of patients.
      - Studies where the subject is anything else, such as a drug, bed, gene, are excluded.
   c. A prediction model must be developed and evaluated.
      - Studies that do not train and evaluate a parameterized model, e.g. only report the odd-ratios of covariates, only run statistical tests, or do not evaluate the developed model, are excluded.
   d. All clinical domains are relevant, such as intensive, radiology, general practitioner, or psychiatric care.
2. **The model predictors must be based on information extracted from unstructured clinical text data in an EHR database.**
   a. The information is extracted from human-readable text data in an EHR database.
      - Studies that only use text data from other sources, such as social media, literature, recordings, transcripts, or genetic data are excluded.
   b. The extracted information is used as covariates in the model.
      - Studies that only use the information to define the outcome or target patient cohorts are excluded.
   c. The model must a least use information from unstructured text data, but it can be used in combination with structured data.
3. **The information from the unstructured clinical text data must be automatically extracted in a data-driven manner.**
   a. The extraction must be data-driven, meaning that the extraction is exploratory and it should not be known beforehand what information from the text data is important for model development, i.e. the extraction must not be driven by mere intuition, personal experience, or existing knowledge.
      - Studies that extract only a limited number or specific set of concepts from text data, for example, a small set of vital signs, the smoking status, or only the diagnosis of diabetes, are excluded.
   b. The extraction must be done automatically.
      - Studies are excluded if the information is manually extracted from the text data.

### **Supplementary tables**

*Table S1. Search strategy*

| **Clause** | **Field** | **Search terms** |
| --- | --- | --- |
| 1 | Topic | 'prediction model' OR 'prediction' |
|  | Title, abstract, keywords | ((predict* OR prognost*) NEAR/3 (model* OR risk* OR rule*)) OR  ((early) NEAR/3 (predict* OR ident* OR risk*)) |
|  | Title | predict* OR prognost* OR  ((risk*) NEAR/6 (model* OR estimat* OR adjust* OR assess* OR identif* OR validat* OR forecast* OR scor* OR factor*)) |
| 2 | Topic | 'medical record'(explode) |
|  | Title, abstract, keywords | EHR OR EHRs OR EMR OR EMRs OR clinical* OR nursing OR nurse* OR physician* OR doctor* OR medical* OR healthcare* OR health-care OR patient* OR  ((electronic* OR medical) NEAR/3 (record* OR data*)) OR  ((patient*) NEAR/3 (record* OR histor* OR data*)) |
| 3 | Topic | 'natural language processing' OR 'text mining' |
|  | Title, abstract, keywords | NLP OR mine OR text_mining OR  ((data*) NEAR/4 (unstruct* or raw)) OR  ((natural) NEAR/3 (language* OR text*)) OR  ((unstruct* OR free OR form OR raw OR embedding) NEAR/4 (text OR textual* OR texts OR note OR notes OR phrase*)) OR  ((clinical* OR nursing OR nurse* OR doctor* OR medical* OR healthcare* OR health-care OR feature* OR patient*) NEAR/4 (text OR textual* OR texts OR note OR notes OR phrase*)) OR  ((text OR word* OR textual* OR texts OR note OR notes OR phrase* OR unstruct* OR linguist* OR semantic*)NEAR/4 (feature* OR predictor* OR covariate*)) OR  (topic near/2 model*) |
| 4 | Publication year | [2005-2020] |
|  | Limit | [English] |
|  | Limit | NOT [conference abstract] |

*Table S2. Characteristics of studies included in the review (n=126)*

| **Study** | **Publication**  **year** | **Country of dataset** | **Clinical care settings** | **Number of Prediction Problems** | **Feature Sets**  **compared** |
| --- | --- | --- | --- | --- | --- |
| Halpern [30] | 2012 | USA | Emergency care | 1 | T |
| Karnik [31] | 2012 | USA | Hospital care | 1 | T:S:C |
| Lehman [32] | 2012 | USA | Hospital care | 1 | T:S:C |
| Li [33] | 2012 | USA | Emergency care | 1 | C |
| Huang [34] | 2014 | USA | Psychiatric or mental health care | 2 | C |
| Huddar [35] | 2014 | USA | Intensive care | 1 | T |
| Kontio [36] | 2014 | Finland | Hospital care | 1 | T:S:C |
| Poulin [37] | 2014 | USA | Psychiatric or mental health care | 1 | T |
| Walsh [38] | 2014 | USA | Hospital care | 1 | T:S:C |
| Caballero [39] | 2015 | USA | Hospital care | 1 | S:C |
| Marafino [40] | 2015 | USA | Intensive care | 1 | T |
| Perotte [41] | 2015 | USA | Hospital care | 1 | T:S:C |
| Roysden [42] | 2015 | USA | Hospital care | 1 | C |
| Cohen [43] | 2016 | USA | Hospital care | 1 | T:C |
| Hassanpour [44] | 2016 | USA | Radiology | 1 | T |
| Hu [45] | 2016 | China | Hospital care | 1 | T |
| Luo [46] | 2016 | USA | Intensive care | 1 | S:C |
| McCoy [47] | 2016 | USA | Hospital care | 1 | S:C |
| Miotto [48] | 2016 | USA | Hospital care | 1 | C |
| Rumshisky [49] | 2016 | USA | Hospital care | 1 | S:C |
| Soguero-Ruiz [50] | 2016 | Norway | Hospital care | 1 | T:S:C |
| Temple [51] | 2016 | USA | Intensive care | 1 | T:S:C |
| Buchan [52] | 2017 | USA | Hospital care | 1 | T |
| Frost [53] | 2017 | Canada | Emergency care | 2 | T |
| Hong [54] | 2017 | South Korea | Hospital care | 1 | C |
| Lucini [55] | 2017 | Brazil | Emergency care | 1 | T |
| Zhang [56] | 2017 | USA | Emergency care | 1 | T:S:C |
| Adamou [57] | 2018 | UK | Psychiatric or mental health care | 1 | S:C |
| Bahl [58] | 2018 | USA | Hospital care | 1 | C |
| Banerjee [59] | 2018 | USA | Hospital care | 1 | T |
| Boag [60] | 2018 | USA | Intensive care | 4 | T |
| Coulet [61] | 2018 | USA | Hospital care | 1 | C |
| Gligorijevic [62] | 2018 | USA | Emergency care | 1 | T:S:C |
| Golas [63] | 2018 | USA | Hospital care | 1 | C |
| Huang [64] | 2018 | USA | Hospital care | 1 | C |
| Ian [65] | 2018 | USA | Intensive care | 1 | S:C |
| Krishnan [66] | 2018 | USA | Intensive care | 1 | T |
| Li [67] | 2018 | USA | Hospital care | 1 | T |
| Marafino [21] | 2018 | USA | Intensive care | 1 | S:C |
| Menger [68] | 2018 | Netherlands | Psychiatric or mental health care | 1 | T |
| Parreco [69] | 2018 | USA | Intensive care | 1 | T:S:C |
| Rajkomar [70] | 2018 | USA | Hospital care | 3 | C |
| Sundararaman [71] | 2018 | USA | Hospital care | 1 | T:S:C |
| Sushil [72] | 2018 | USA | Intensive care | 2 | T |
| Weissman [73] | 2018 | USA | Intensive care | 1 | S:C |
| Yang [74] | 2018 | China | Hospital care | 1 | T |
| Afshar [75] | 2019 | USA | Hospital care | 1 | T |
| Akbilgic [76] | 2019 | USA | Surgery care | 1 | T:S:C |
| Alvarez-Mellado [77] | 2019 | USA | Psychiatric or mental health care | 1 | S:C |
| Apostolova [78] | 2019 | USA | Hospital care | 1 | S:C |
| Beeksma [79] | 2019 | Netherlands | Outpatient care | 1 | S:C |
| Brown [80] | 2019 | USA | Surgery care | 1 | T |
| Chen [81] | 2019 | USA | Intensive care | 1 | T:C |
| da Silva [82] | 2019 | Brazil | Surgery care | 1 | T |
| Danielsen [83] | 2019 | Denmark | Psychiatric or mental health care | 1 | C |
| Danilov [84] | 2019 | Russia | Surgery care | 1 | T |
| Gong [85] | 2019 | China | Hospital care | 1 | C |
| Khadanga [86] | 2019 | USA | Intensive care | 2 | T:S:C |
| Kongburan [87] | 2019 | USA | Intensive care | 1 | S:C |
| Korach [88] | 2019 | USA | Hospital care | 1 | T:S:C |
| Krishnan [89] | 2019 | USA | Intensive care | 1 | T |
| Liu [90] | 2019 | USA | Intensive care | 1 | S:C |
| Mahajan [91] | 2019 | USA | Hospital care | 1 | T:S:C |
| Makino [92] | 2019 | Japan | Hospital care | 1 | S:C |
| Menger [22] | 2019 | Netherlands | Psychiatric or mental health care | 1 | T |
| Nakayama [93] | 2019 | USA | Hospital care | 1 | S:C |
| Payrovnaziri [94] | 2019 | USA | Hospital care | 1 | S:C |
| Ross [95] | 2019 | USA | Hospital care | 1 | S:C |
| Shin [96] | 2019 | USA | Hospital care | 1 | C |
| Si [97] | 2019 | USA | Hospital care | 1 | T |
| Sterling [98] | 2019 | USA | Emergency care | 1 | T |
| Sun [99] | 2019 | USA | Intensive care | 1 | T:S:C |
| Wang [100] | 2019 | USA | Hospital care | 1 | C |
| Weissman [101] | 2019 | USA | Hospital care | 1 | T |
| Yang [102] | 2019 | China | Hospital care | 1 | T |
| Zhang [103] | 2019 | USA | Emergency care | 1 | T:S:C |
| Bacchi [104] | 2020 | Australia | Emergency care | 2 | T |
| Barash [105] | 2020 | Israel | Intensive care | 1 | T:S:C |
| Barber [106] | 2020 | USA | Surgery care | 2 | T:S:C |
| Baxter [107] | 2020 | USA | Hospital care | 1 | T:S |
| Ben Miled [108] | 2020 | USA | unclear | 1 | T:C |
| Chen [109] | 2020 | China | Surgery care | 1 | S:C |
| Chen [110] | 2020 | China | Surgery care | 1 | T:S:C |
| Chen [111] | 2020 | Taiwan | Emergency care | 1 | S:C |
| Chen [112] | 2020 | Taiwan | Emergency care | 1 | S:C |
| Danilov [113] | 2020 | Russia | Surgery care | 1 | T |
| Fernandes [114] | 2020 | Portugal, USA | Emergency care | 2 | C |
| Fernandes [115] | 2020 | Portugal | Emergency care | 1 | S:C |
| Gensheimer [116] | 2020 | USA | Hospital care | 1 | C |
| Goodwin [117] | 2020 | USA | Hospital care | 3 | T |
| Guo [118] | 2020 | USA | Hospital care | 1 | C |
| Hane [119] | 2020 | USA | Claims | 1 | S:C |
| Hashir [120] | 2020 | USA | Hospital care | 1 | T:S:C |
| Heo [121] | 2020 | South-Korea | Hospital care | 1 | T |
| Hsu [122] | 2020 | USA | Hospital care | 2 | T:S:C |
| Izquierdo [123] | 2020 | Spain | Hospital care | 1 | T |
| Korach [124] | 2020 | USA | Hospital care | 1 | T |
| Le [125] | 2020 | USA | Intensive care | 1 | S:C |
| Lee [126] | 2020 | USA | Outpatient care | 2 | C |
| Levis [127] | 2020 | USA | Psychiatric or mental health care | 1 | T |
| Li [128] | 2020 | USA | Intensive care | 1 | C |
| Meng [129] | 2020 | not reported | unclear | 1 | S:C |
| Mohammadi [130] | 2020 | USA | Surgery care | 2 | T:S |
| Mugisha [131] | 2020 | USA | Intensive care | 1 | T:S:C |
| Nakatani [132] | 2020 | Japan | Hospital care | 1 | T |
| Obeid [133] | 2020 | USA | Hospital care | 1 | T |
| Roquette [134] | 2020 | Brazil | Hospital care | 1 | S:C |
| Shukla [135] | 2020 | USA | Hospital care | 1 | T:S:C |
| Sterckx [136] | 2020 | Belgium | Hospital care | 1 | T:S:C |
| Sterling [137] | 2020 | USA | Emergency care | 1 | C |
| Tahayori [138] | 2020 | Australia | Emergency care | 1 | T |
| Topaz [139] | 2020 | USA | Outpatient care | 1 | T |
| Wang [140] | 2020 | USA | Hospital care | 1 | T:C |
| Weegar [141] | 2020 | Sweden | Hospital care | 1 | T:S:C |
| Xu [142] | 2020 | USA | Hospital care | 1 | T |
| Ye [143] | 2020 | USA | Hospital care | 1 | T:S |
| Zhang [144] | 2020 | USA | Hospital care | 3 | T:S:C |
| Boag [145] | 2021 | USA | Psychiatric or mental health care | 1 | T:S:C |
| Chen [146] | 2021 | Taiwan | Emergency care | 1 | C |
| Goh [147] | 2021 | Singapore | Hospital care | 1 | S:C |
| Klang [148] | 2021 | USA | Emergency care | 1 | T:S:C |
| Muhlestein [149] | 2021 | USA | Surgery care | 1 | T:S |
| Oliwa [150] | 2021 | USA | Outpatient care | 1 | T |
| Ribelles [151] | 2021 | Spain | Hospital care | 1 | T:S |
| Tang [152] | 2021 | USA | Hospital care | 1 | T |
| Yang [153] | 2021 | USA | Hospital care | 1 | T:C |

### **Supplementary figures**

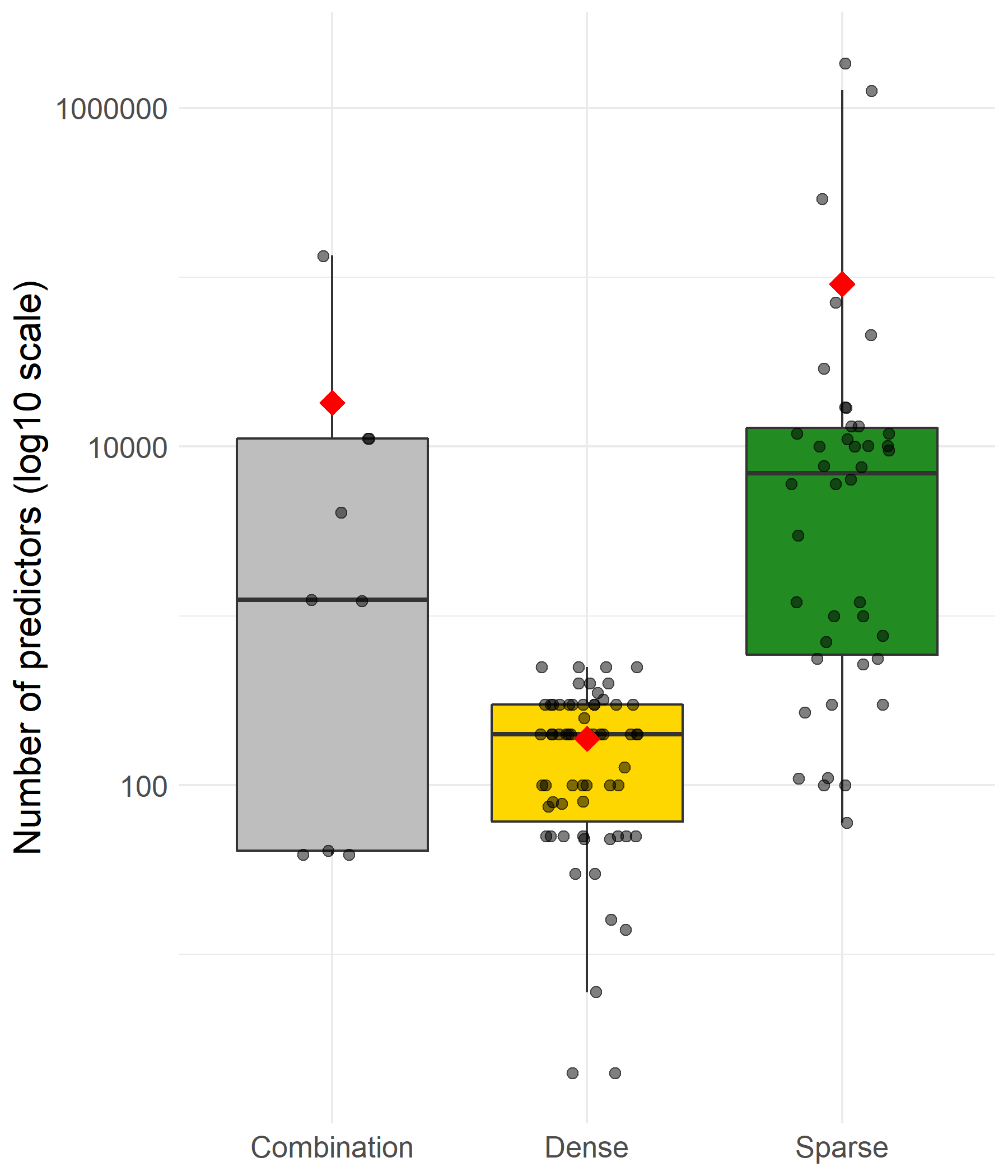

*Figure S1. Number of predictors in combined, dense, or sparse text representations. The mean is indicated by the red diamond and the points represent the underlying data.*

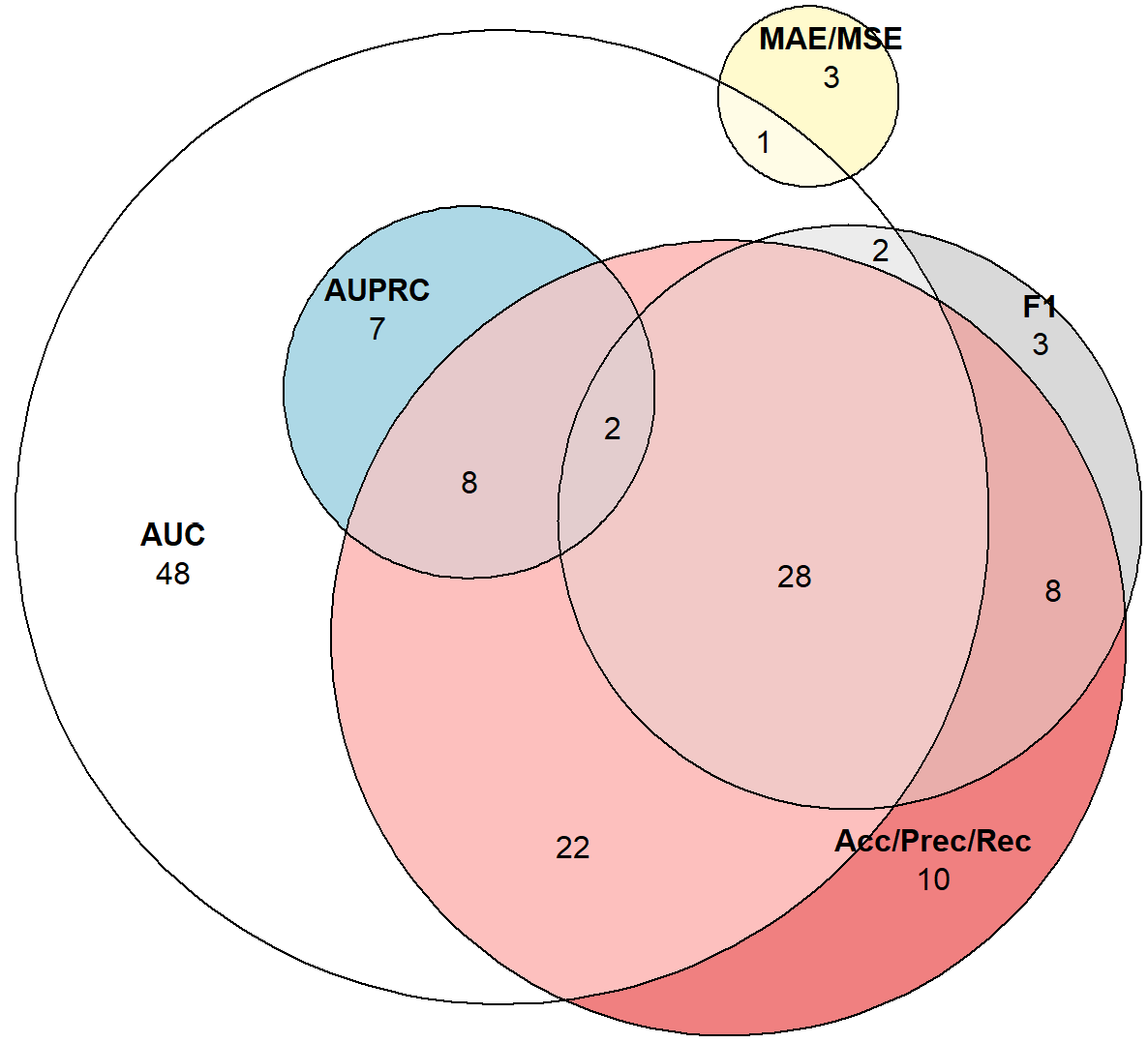

*Figure S2. Euler diagram depicting the combined reporting of different performance metrics in the 145 prediction problems. AUC: Area under the receiver operator curve, AUPRC: area under the precision-recall curve, MAE: Mean absolute error, MSE: mean squared error, F1: F1 score, Acc/Prec/Rec: reporting of accuracy, sensitivity (or recall), specificity, or positive predictive value (or precision).*
